# Transforming Primary Healthcare (PHC) for Adiposity-Based Chronic Disease (ABCD) and Cardiovascular-Kidney-Metabolic (CKM) Syndrome: Integrating Endothelin-1 (ET-1) and TG/HDL-C Ratio for Subclinical Cardiovascular Risk Stratification

**DOI:** 10.1101/2025.06.11.25329411

**Authors:** Aldian Arie Pratama, Agustin Iskandar, Singgih Pudjo Wahono

## Abstract

**Objective:** To fill the gaps in primary healthcare (PHC) service delivery strategies focused on adiposity-based chronic disease and chronic kidney metabolic syndrome in the non-communicable diseases (NCD) context, focus on healthcare transformation and practice redesign.

**Design:** an experimental analytics study in tertiary care hospitals with a study population of adults aged 30-65 years with ABCD (BMI ≥ 25 kg/m^2^) or stage 1 CKM syndrome (defined by coexisting overweight and/or obesity conditions). The study is designed to address locally relevant health priorities, specifically, the rising burden of cardiovascular-renal-metabolic (CKM) diseases and the need for early, affordable risk stratification tools such as ET-1 and the TG/HDL-C ratio. The research aims to generate evidence that can directly inform local clinical practice and health policy, thereby benefiting both study participants and the broader community. The collected data were then analyzed statistically; the quantification method used body mass index (BMI), then examination of ET-1 levels was carried out using the ELISA method, and lipid fractions using the enzymatic method.

**Setting:** The study is designed to address locally relevant health priorities, specifically, the rising burden of cardiovascular-renal-metabolic (CKM) diseases and the need for early, affordable risk stratification tools such as ET-1 and the TG/HDL-C ratio. The research aims to generate evidence that can directly inform local clinical practice and health policy, thereby benefiting both study participants and the broader community. The study included adults aged 30 to 65 years, consistent with common age ranges for cardiovascular risk studies in LMICs, diagnosed with ABCD or stage I CKM syndrome, including obesity (BMI ≥ 25 kg/m^2^), and 97 participants met the inclusion criteria, with the rest of the subjects excluded.

**Participants:** The study included adults aged 30 to 65 years, consistent with common age ranges for cardiovascular risk studies in LMICs, diagnosed with ABCD or stage I CKM syndrome, including obesity (BMI ≥ 25 kg/m^2^), and 97 participants met the inclusion criteria, with the rest of the subjects excluded.

**Results:** The collected data were then analyzed statistically with distribution tests, difference tests, correlation tests, and multivariate analysis. The difference test of ET-1 levels and TG/HDL-C ratios to the degree of obesity using the one-way ANOVA test found significant differences in ET-1 levels and TG/HDL-C ratios to the degree of obesity (p-value < 0.001). and (p-value < 0.001). Where based on the least significant difference in the non-obese sub-population against obesity (p-value 0.051) and significantly different from the obesity II population (p-value < 0.001), then the LSD test of the TG/HDL-C ratio against the degree of obesity was significantly different in the non-obese population against obesity I (p-value 0.002) and non-obese against obesity II (p-value < 0.001). From the multivariate analysis, there were statistically significant differences in the mean values of the ET-1 variable between the obese II sub-population and the non-obese sub-population OR: 216.29 (95% CI: 91.25 to 341.33; p-value 0.000), as well as between obesity II and obesity I OR: 119.49 (95% CI: 60.68 to 178.29; p-value 0.000). Meanwhile, the TG/HDL-C ratio variable had a statistically significant effect on the non-obese, obesity I, and obesity II populations OR: 3.16 (95% CI: 0.71 to 5.52; p-value < 0.001).

From this study, all subjects were indicated to have endothelial dysfunction, where, based on the TG/HDL-C ratio, all subjects could be classified as having insulin resistance, and, based on the atherogenic index of plasma (AIP) algorithm, the study population was stratified into moderate risk for first-time incidence of atherosclerotic cardiovascular disease (n=10) and high risk for first-time incidence of atherosclerotic cardiovascular disease (n=87).

**Conclusion:** Integrating plasma endothelin-1 (ET-1) levels and the triglyceride-to-HDL cholesterol (TG/HDL-C) ratio into cardiovascular risk assessment frameworks offers a promising strategy to enhance early detection and management of adiposity-based chronic disease (ABCD) and cardiovascular-kidney-metabolic (CKM) syndrome, particularly in low- and middle-income countries (LMICs). These biomarkers reflect key pathophysiological processes, endothelial dysfunction and atherogenic dyslipidemia, that underpin subclinical cardiovascular and renal injury. Their combined application in primary healthcare settings can bridge critical gaps in current non-communicable disease (NCD) care by enabling precision risk stratification, guiding timely interventions, and ultimately reducing morbidity and mortality.

Future large-scale, longitudinal studies are warranted to validate these findings and support guideline incorporation, thereby advancing healthcare transformation aligned with national health security (NHS) and sustainable development goals (SDGs).

## INTRODUCTION

The global burden of non-communicable diseases (NCDs), particularly adiposity-based chronic disease (ABCD) and cardiovascular-kidney-metabolic (CKM) syndrome, continues to escalate, posing significant challenges to healthcare systems worldwide, especially in low- and middle-income countries (LMICs). ABCD, characterized by excess adiposity and its metabolic consequences, is intricately linked with CKM syndrome, a cluster of interrelated conditions that markedly increase the risk of cardiovascular disease (CVD) and chronic kidney disease (CKD). ^1^

It is well recognized that non-communicable diseases (NCDs) represent the leading cause of mortality globally, accounting for more than 73% of deaths each year. In 2010, nearly 80% of these deaths occurred in low- and middle-income countries (LMICs), regions undergoing rapid demographic shifts, including population aging, urbanization, increased tobacco consumption, and evolving patterns of diet and obesity. However, the primary healthcare (PHC) systems in LMICs—traditionally focused on infectious diseases and maternal-child health—remain inadequately equipped to manage the growing burden of NCDs. ^2,3^

Yet the primary healthcare (PHC) systems of LMICs, historically oriented to infectious diseases and maternal and child health, are not well designed to integrate NCD care. ^4^

Despite advances in understanding the pathophysiology of ABCD and CKM, current primary healthcare (PHC) strategies often lack effective tools for early risk stratification, limiting timely intervention. Emerging evidence highlights the potential of integrating biomarkers such as endothelin-1 (ET-1), a potent vasoconstrictor implicated in endothelial dysfunction and renal injury, and the triglyceride-to-HDL cholesterol (TG/HDL-C) ratio, a validated surrogate marker of atherogenic dyslipidemia, as complementary indicators of subclinical cardiovascular risk. ^5^

ET-1 levels have been shown to correlate with declining glomerular filtration rate and cardiovascular events, while the TG/HDL-C ratio predicts insulin resistance and coronary artery disease even in normolipidemic individuals. However, the application of these biomarkers in PHC settings, particularly in resource-limited environments, remains underexplored. There is a critical gap in current clinical guidelines regarding their use for early detection and risk stratification in ABCD and CKM populations. Addressing this gap aligns with global health priorities, including the sustainable development goals (SDGs) and national health security frameworks, by promoting precision medicine approaches that are both cost-effective and scalable. The main aim of this study is to propose a transformative PHC model that integrates ET-1 and TG/HDL-C ratio measurements to enhance subclinical cardiovascular risk assessment in individuals with ABCD and CKM syndromes. ^6^

This approach seeks to facilitate earlier diagnosis, guide personalized interventions, and ultimately improve cardiovascular and renal outcomes in high-risk populations. ^7^

Incorporating these biomarkers within a predictive, preventive, and personalized medicine (3PM) framework offers a transformative opportunity to shift from reactive to proactive healthcare. The predictive aspects enable identification of individuals at heightened risk before clinical manifestations, while preventive strategies can be tailored based on biomarker profiles to mitigate disease progression. Personalized interventions, informed by ET-1 and the TG/HDL-C ratio, facilitate optimized therapeutic decisions, improving efficacy and minimizing adverse effects. However, the application of these biomarkers in PHC settings, particularly in resource-limited environments, remains underexplored. There is a critical gap in current clinical guidelines.

Regarding their use for early detection and risk stratification in ABCD and CKM populations. Addressing this gap aligns with global health priorities, including the Sustainable Development Goals (SDGs) and national health security (NHS) frameworks, by promoting precision medicine approaches that are both cost-effective and scalable. The main aim of this study is to propose a transformative PHC model that integrates ET-1 and TG/HDL-C ratio measurements within a 3PM approach to enhance subclinical cardiovascular risk assessment in individuals with ABCD and CKM syndrome.

This approach facilitates early identification of at-risk individuals (predictive), enables tailored interventions to halt or slow disease progression (preventive), and supports individualized management strategies (personalized), thereby improving clinical outcomes and resource utilization. Moreover, embedding this 3PM approach into PHC directly supports national health security (NHS) frameworks and the United Nations Sustainable Development Goals (SDGs).

NHS initiatives emphasize strengthening health systems to manage NCDs effectively, ensuring equitable access to diagnostics and care, and enhancing population resilience against chronic diseases. Concurrently, SDGs, particularly Goal 3, which aims to ensure healthy lives and promote well-being for all at all ages, prioritize reducing premature mortality from NCDs through prevention and treatment. The World Health Organization and national policies advocate for integrating cost-effective, scalable interventions into PHC to achieve these targets, highlighting the importance of innovative biomarker-driven strategies in resource-limited settings. ^8^

The primary objective of this study is to evaluate the effectiveness of integrating plasma endothelin-1 (ET-1) levels and triglyceride-to-HDL cholesterol (the TG/HDL-C) ratio within a predictive, preventive, and personalized medicine (3PM) framework to enhance subclinical cardiovascular risk stratification in individuals with adiposity-based chronic disease (ABCD) and cardiovascular-kidney-metabolic (CKM) syndrome in primary healthcare settings.

### Specific objectives include

1. To assess the association between ET-1 levels and the TG/HDL-C ratio with markers of subclinical cardiovascular and renal dysfunction in ABCD and CKM populations.
2. To determine the predictive value of combined ET-1 and the TG/HDL-C measurements for early identification of individuals at high risk of cardiovascular events and renal progression.
3. To explore the feasibility and clinical utility of implementing ET-1 and TG/HDL-C ratio testing in resource-limited primary healthcare environments.
4. To align the biomarker-based risk stratification approach with national health security (NHS) goals and Sustainable Development Goals (SDGs) for improving non-communicable disease management and outcomes.

## METHODS

### STUDY DESIGN

This experimental research design of this study employed an analytical cross-sectional design based on the principles of Predictive, Preventive, and Personalized Medicine (3PM). Aiming to evaluate the utility of endothelin-1 (ET-1) and the triglyceride-to-HDL cholesterol (TG/HDL-C) ratio for subclinical cardiovascular risk stratification in individuals with adiposity-based chronic disease (ABCD) and cardiovascular-kidney-metabolic (CKM) syndrome. Participants aged 30-65 years diagnosed with ABCD or CKM syndrome will be recruited in a low- and middle-income country (LMIC) setting, reflecting real-world clinical practice constraints and population diversity. Data collection and measurements: at baseline, participants will undergo comprehensive clinical evaluation, including anthropometric measurements and collection of blood samples for ET-1 quantification (via ELISA) and lipid profiling to calculate the TG/HDL-C ratio. Additional data on demographics, medical history, lifestyle factors, and medication use will be collected. The study will analyze the prediction capacity of ET-1 and the TG/HDL-C ratio individually and in combination. The study embraces 3PM by focusing on early prediction, targeted prevention strategies, and personalized risk profiling. Ethical considerations: The study protocol will adhere to international ethical standards and receive approval from relevant institutional review boards. Informed consent will be obtained from all participants, ensuring confidentiality and data protection.

### SETTING

This study will be conducted in primary healthcare centers and tertiary hospitals located in urban areas of a low- and middle-income country (LMIC), reflecting the healthcare delivery context where adiposity-based chronic disease (ABCD) and cardiovascular-kidney-metabolic (CKM) syndrome are highly prevalent but often underdiagnosed. The selected sites represent typical resource-constrained environments characterized by limited access to advanced diagnostic tools, workforce shortages, and diverse patient populations with varying socioeconomic backgrounds, including routine cardiovascular and metabolic disease management, enabling integration of biomarker assessments such as endothelin-1 (ET-1) and triglyceride-to-HDL cholesterol (TG/HDL-C) ratio into existing clinical workflows. Laboratory analysis will be performed using validated, cost-effective assays compatible with local infrastructure to ensure feasibility and sustainability. This setting allows for evaluation of the predictive, preventive, and personalized medicine (3PM) approach in real-world conditions aligned with national health security priorities and Sustainable Development Goals (SDGs), aiming to improve early detection and management of subclinical cardiovascular risk in ABCD and CKM patients within PHC frameworks.

### CONCEPTUAL FRAMEWORK

This study’s conceptual framework centers on the paradoxical role of adiposity in cardiovascular-renal-metabolic (CVRM) syndrome, integrating key biomarkers—endothelin-1 (ET-1) and the triglyceride-to-HDL cholesterol (TG/HDL-C) ratio—to improve subclinical cardiovascular risk stratification within a predictive, preventive, and personalized medicine (3PM) approach. (Figure 1). ^11^

**Figure 1.**
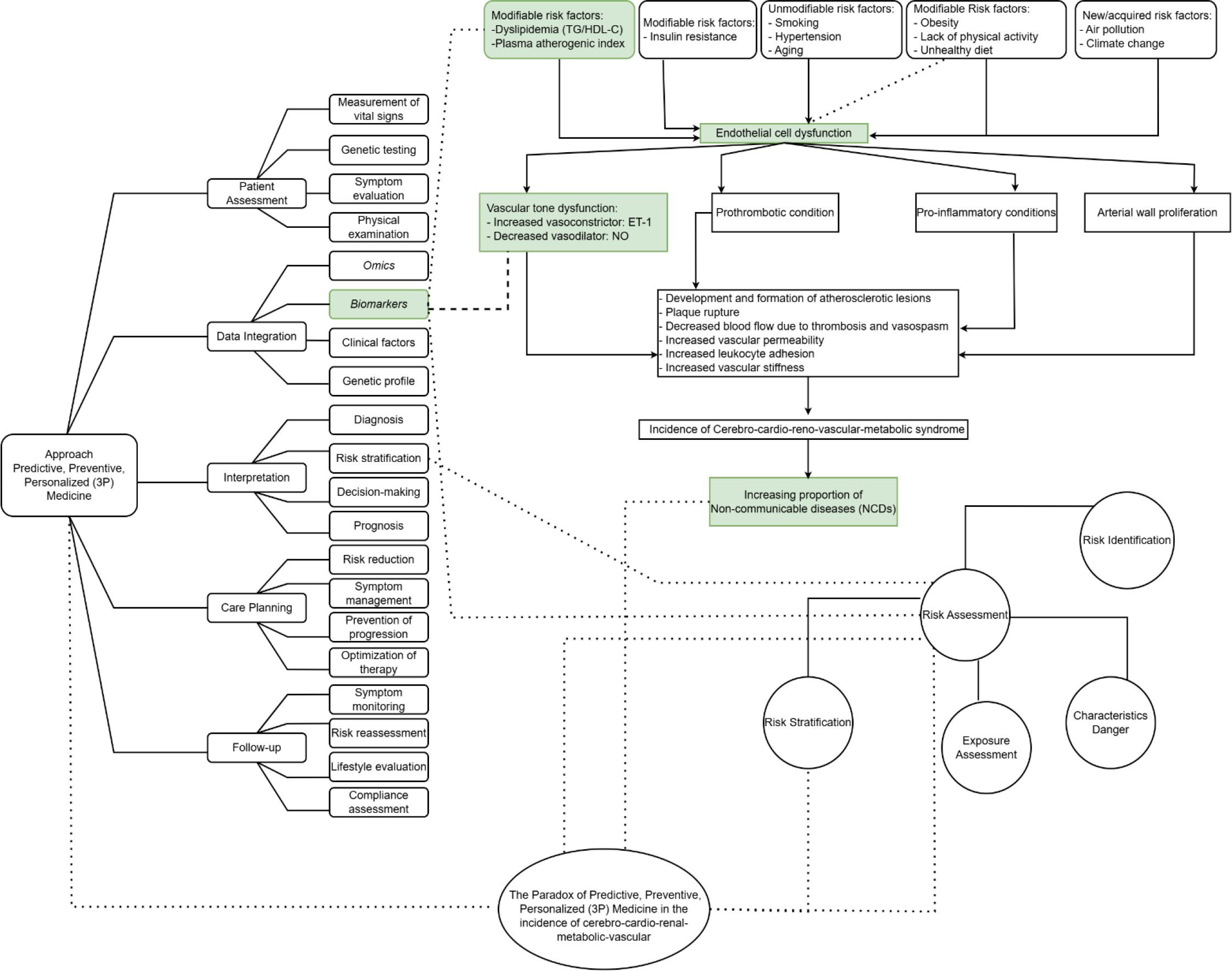
The conceptualization of Predictive, Preventive, Personalized Medicine (3PM) with paradoxical adiposity-based chronic disease (ABCD) and cardiovascular-kidney-metabolic (CKM) syndrome with an increased proportion of non-communicable diseases (NCDs). ^11^

**Figure 2.**
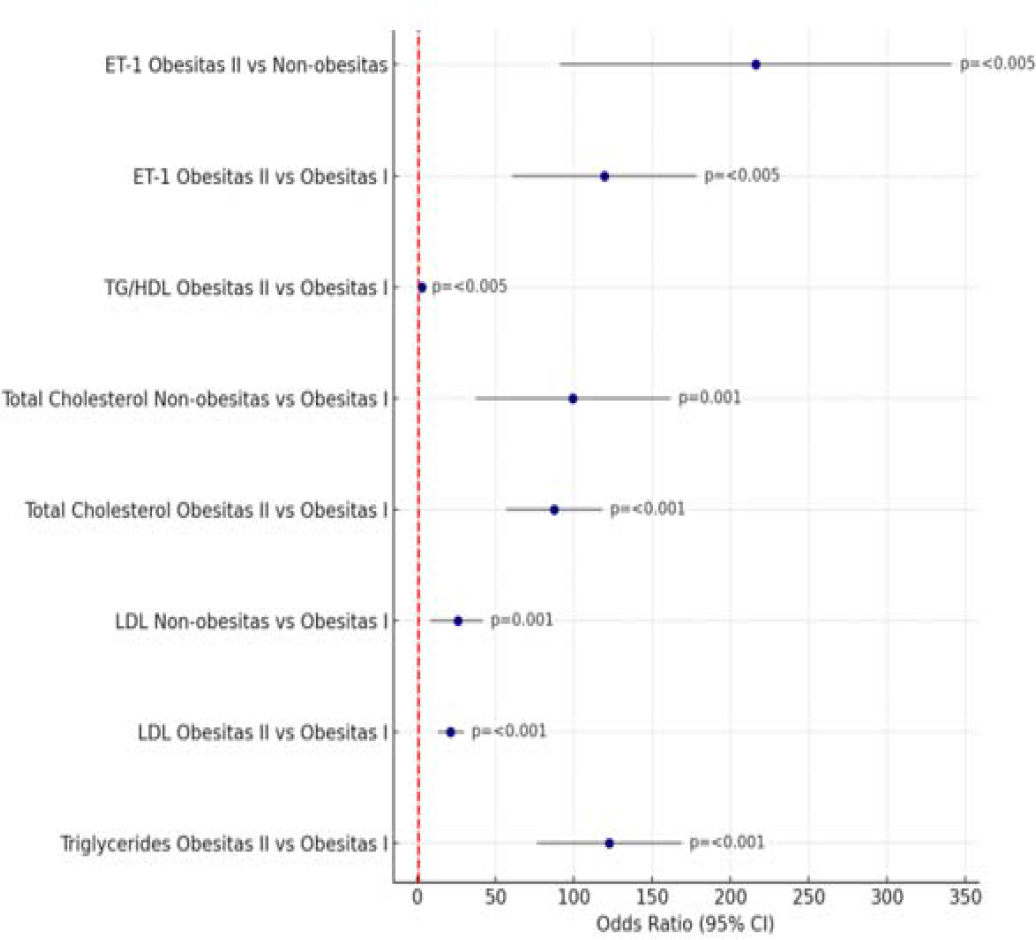
Forest plot of the results of the multivariate analysis of ET-1 and lipid fractions on the degree of obesity^12^

With key components: paradoxical obesity in CVRM syndrome Although obesity is a well-established risk factor for cardiovascular disease (CVD) and chronic kidney disease (CKD), paradoxical obesity refers to observations where increased adiposity sometimes associates with unexpected protective cardiovascular outcomes or variable risk profiles. ^12^ This paradox complicates risk assessment and clinical decision-making in adiposity-based chronic disease (ABCD) and CKM syndrome. ^13,14^ Pathophysiological mediator endothelin-1 (ET-1): a potent vasoconstrictor and pro-inflammatory peptide elevated in endothelial dysfunction and renal injury. ET-1 levels rise with worsening renal function and contribute to vascular remodeling and hypertension, linking adiposity to cardiovascular and renal pathology. ^15^ TG/HDL-C ratio: reflects atherogenic dyslipidemia and insulin resistance, serving as a surrogate marker for lipid-related cardiovascular risk. Elevated TG/HDL-C ratios correlate with increased plaque vulnerability and adverse cardiovascular events. ^12^

Biomarker interaction and paradox resolution The interplay between ET-1 and the TG/HDL-C ratio may elucidate the mechanism underlying paradoxical obesity by differentiating metabolically healthy versus unhealthy adiposity phenotypes and identifying subclinical vascular damage not apparent through traditional measures such as BMI alone. ^16^ Predictive, Preventive, and Personalized Medicine (3PM) integration Predictive: Early detection of elevated ET-1 and TG/HDL-C ratio enables identification of individuals at heightened risk despite paradoxical obesity presentations. Preventive: Targeted interventions can be implemented to modulate ET-1 pathways and lipid abnormalities, preventing progression to overt CVD and CKD. Personalized: Risk stratification based on combined biomarker profiles supports individualized management plans tailored to patient-specific pathophysiology. Health system context: Embedding this biomarker-driven 3PM model within primary healthcare aligns with national health security objectives and Sustainable Development Goals (SDGs), promoting equitable, cost-effective strategies for managing ABCD and CKM syndromes in low- and middle-income countries. ^13^**Open access**

## RESULTS

A total of 97 participants were diagnosed with adiposity-based chronic disease (ABCD) and cardiovascular-kidney-metabolic (CKM) syndrome. Baseline characteristics are shown in Table 1-4^**12**^

**Table 1.** Characteristics of this study^**12**^

| Characteristics | Non-obesity (n=5) | Obesity I (n=27) | Obesity II (n=65) | p-value* |
| --- | --- | --- | --- | --- |
|  | Mean ± SD | Mean ± SD | Mean ± SD |  |
| Age (years) | 43,4±5,89 | 37,89±8,38 | 39,96±8,05 | 0,642 |
| BMI (kg/m<sup>2</sup>) | 24,06±0,61 | 34,43±2,73 | 27,62±1,08 | 0,533 |
| ET-1 | 289,40±100,83 | 386,21±104,69 | 505,70±107,66 | <0,001* |
| Triglycerides | 172,47±71,55 | 295,07±98,54 | 307,2±130,37 | <0,001* |
| Total-Cholesterol | 193,15±61,34 | 280,55±39,0 | 292,6±31,18 | 0,103 |
| LDL-Cholesterol | 136,3±16,81 | 157,67±9,39 | 161,5±9,45 | 0,091 |
| HDL-Cholesterol | 50,15±5,40 | 48,4±6,02 | 46,81±5,51 | 0,303 |
| TG/HDL ratio | 3,56±1,94 | 6,50±2,45 | 6,67±3,48 | <0,001* |

\*significant if p-value <0,05

**Table 2.** Research difference test ^12^

| Variables | n (%) | Non-obesity<br>(Mean±SD) | Obesity I<br>(Mean±SD) | Obesity II<br>(Mean±SD) | p-value* |
| --- | --- | --- | --- | --- | --- |
| ET-1 | 97(100%) | 289,40 ± 100,83 | 386,21 ± 104,69 | 505,70 ± 107,66 | <0,001* |
| TG/HDL | 97(100%) | 3,56±1,94 | 6,50±2,45 | 6,67±3,48 | <0,001* |

\*one-way Anova significant if p-value<0,005

**Table 3.** Post hoc test in this study^12^

| Variables | Sub-population study |  | p-value* |
| --- | --- | --- | --- |
| ET-1 | Non-obesity | Obesity I | <0,001* |
|  |  | Obesity II | <0,001* |
|  | Obesity I | Obesity II | 0,051 |
| TG/HDL ratio | Non-obesity | Obesity I | <0,001* |
|  |  | Obesity II | <0,001* |
|  | Obesity I | Obesity II | 0,002* |

\*significant if p-value<0,005

**Table 4.** Multivariate analysis of ET-1 and TG/HDL-C ratio against obesity degree ^12^

| Variables | BMI categorize | Sub-categorize BMI | p-value* | Odds Ratio (OR) (95% CI) |
| --- | --- | --- | --- | --- |
| ET-1 | Non-obesity | Obesity I | 0,152 | -96,80 (-215,99 s/d 22,39) |
|  |  | Obesity II | 0,000* | -216,29* (-341,33 s/d -91,25) |
|  | Obesity I | Non-obesity | 0,152 | 96,80 (-22,39 s/d 215,99) |
|  |  | Obesity II | 0,000* | -119,49* (-2,42 s/d 6,58) |
|  | Obesity II | Non-obesity | 0,000* | 216,29* (91,25 s/d 341,33) |
|  |  | Obesity I | 0,000* | 119,49* (60,87 s/d 178,29) |
| TG/HDL | Non-obesity | Obesity I | 0,006 | 3,11* (0,71 s/d 5,15) |
|  |  | Obesity II | 1,000 | 0,17 (-2,34 s/d 2,69) |
|  | Obesity I | Non-obesity | 0,006 | -3,11* (-5,51 s/d -0,71) |
|  |  | Obesity II | 0,000* | -2,94* (-4,12 s/d -1,75) |
|  | Obesity II | Non-obesity | 1,000 | -0,17 (-2,69 s/d 2,34) |
|  |  | Obesity I | 0,000* | 2,94* (1,75 s/d 4,12) |

\*significant if p-value<0,005

In this study, the subjects collected (n=97), where the population that met the inclusion criteria were non-obesity (n=5), obesity (n=27), and obesity II (n=65).

The ET-1 difference test in non-obese and obese individuals was conducted using one-way ANOVA, because the data distribution was normal when the data normality test was conducted using the Kolmogorov-Smirnov test.

The results of the ET-1 difference test and the TG/HDL ratio against the degree of obesity using the one-way ANOVA method obtained a p-value of <0.001, where the value was significantly different in the ET-1 level variable (p-value <0.005) against the degree of obesity and significantly different in the TG/HDL-C ratio variable (p-value <0.005) against the degree of obesity. So that the Least Significance Difference (LSD) test was carried out to determine the significance in each population group.

According to the post hoc test, the ET-1 level against the degree of obesity was known in the non-obesity population group to be significantly different from obesity I (p-value = <0.001) and significantly different from obesity population group II (p-value = <0.001), whereas in obesity group I it was known not to be significantly different from obesity II (p-value = 0.051).

The TG/HDL-C ratio according to the post hoc test level against the degree of obesity was known in the non-obesity population group to be significantly different from the obesity population group I (p-value= <0.001) and significantly different from the obesity population group II (p-value= <0.001), where obesity I was not significantly different from obesity II (p-value= 0.051).

And according to multivariate analysis, the results are revealed in Table 4.

The results of the multivariate analysis test showed that the results of the ET-1 variable on obesity II had an effect on the non-obesity population (OR = 216.29; 95% CI: 91.25 to 341.33; p-value = 0.000) and on the obesity I population (OR = 119.49; 95% CI: 60.68 to 178.29; p-value = 0.000). The TG/HDL ratio variable for the obesity II population was known to have an effect on the obesity I population (OR = 2.94; 95% CI: 1.75 to 4.12; p-value = 0.000) and reached statistical significance, indicating an increase in the TG/HDL-C ratio as a marker of increased metabolic risk to the degree of obesity.

Interpretation of the forest plot and multivariate analysis results reveals critical insights into the association of endothelin-1 (ET-1) and the TG/HDL-C ratio with obesity severity, highlighting their roles as metabolic risk markers in adiposity-based chronic disease (ABCD) and cardiovascular-kidney-metabolic (CKM) syndrome.

ET-1 and obesity II versus non-obesity: OR = 216.29 (95% CI: 91.25-341.33; p-value = 0.000) This extraordinarily high odds ratio (OR) suggests that elevated ET-1 levels are strongly predictive of severe obesity (obesity II) compared to non-obese individuals. The wide confidence interval (CI) indicates substantial variability in the effect size, but the statistically significant p-value reinforces the robustness of this association.

Biological rationale: ET-1, a potent vasoconstrictor and pro-inflammatory mediator, may drive endothelial dysfunction and insulin resistance, exacerbating adiposity and metabolic dysregulation. Non-obesity vs. obesity I: OR = 119.49 (95% CI: 60.68-178.29; p-value = 0.000) ET-1 levels also show a strong, dose-dependent association with moderate obesity (Obesity I), though the effect size is smaller than for Obesity II. This gradient supports ET-1’s role in adiposity progression.

The TG/HDL-C ratio and obesity severity: obesity I vs. obesity II: OR = 2.94 (95% CI: 1.75-4.12; p-value = 0.000) The TG/HDL-C ratio is significantly higher in obesity II compared to obesity I, with a nearly 3-fold increase in odds per unit rise in the ratio. Clinical implication: this ratio, a marker of atherogenic dyslipidemia and insulin resistance, escalates with worsening obesity, reflecting its utility in stratifying metabolic risk.

Key takeaways: ET-1 as a potent predictor of obesity severity: The exceptionally high OR for ET-1 suggests it is a dominant biomarker for distinguishing obesity categories, potentially reflecting its role in vascular and metabolic dysfunction. However, the extreme effect sizes warrant caution; possible explanations include collinearity: ET-1 may correlate strongly with unmeasured confounders (e.g., visceral adiposity or renal impairment).

Threshold effect: ET-1 levels might exhibit a non-linear relationship, where only values above a critical threshold drive obesity progression. TG/HDL-C ratio as a graded metabolic risk marker: the ratio’s progressive increase across obesity categories aligns with its established role in predicting cardiovascular risk.

The moderate OR (2.94) underscores its relevance in clinical practice for early metabolic risk assessment. Biological synergy: the combined elevation of ET-1 and TG/HDL-C ratio may synergistically amplify oxidative stress, inflammation, and endothelial damage, accelerating ABCD/CKM progression.

Limitations and Recommendations Causality vs. Association: The observational design precludes causal inferences; longitudinal studies are needed to confirm ET-1’s role in obesity pathogenesis. Generalizability: extreme ORs for ET-1 may reflect cohort-specific characteristics (e.g., genetic or environmental factors).

Replication in diverse populations is critical. Clinical translation: while these biomarkers show promise, cost-effectiveness and feasibility of routine ET-1 testing in primary care require evaluation. Conclusion: ET-1 and the TG/HDL-C ratio are robust, complementary biomarkers for obesity-related metabolic risk stratification.

Their integration into primary healthcare protocols could enhance early detection of high-risk ABCD/CKM patients, enabling personalized interventions to mitigate cardiovascular and renal complications. Further mechanistic studies are needed to unravel ET-1’s paradoxical role in adiposity and metabolic dysregulation.

**Graph 1.**
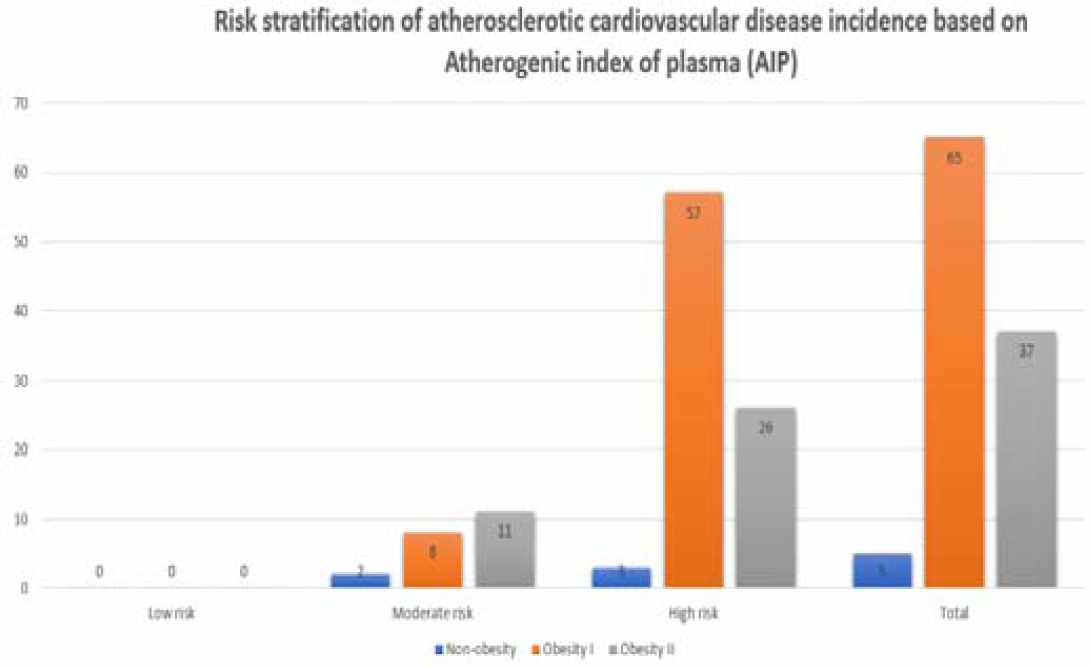
Risk stratification in this study ^12^.

The predictive number of AIP is said to be higher for the incidence of atherosclerosis, where evidence-based medicine (EBM) mentions the cut-off classification of AIP, namely, 0.3-0.11 as low risk, 0.11-0.21 as moderate risk, and >0.21 as high risk for the occurrence of first-time atherosclerotic heart disease.

According to the cut-off from the EBM, the risk stratification of atherosclerotic cardiovascular disease incidence based on AIP, as attached in Graph 1, shows that based on the atherogenic index of plasma (AIP), the population in this study is classified into moderate risk and high risk for first-time heart disease incidence and requires more attention from the perspective of predictive, preventive, and precision medicine (3PM).

## DISCUSSION

### Summary of key findings

The multivariate analysis and corresponding forest plot provide compelling evidence that endothelin-1 (ET-1) and the triglyceride-to-HDL cholesterol (TG/HDL-C) ratio are significant and independent predictors of obesity severity and metabolic risk within the context of adiposity-based chronic disease (ABCD) and cardiovascular-kidney-metabolic (CKM) syndrome. ^26,27,28^ The markedly elevated odds ratios (ORs) for ET-1 in both Obesity II (OR = 216.29; 95% CI: 91.25-341.33) and Obesity I (OR = 119.49; 95% CI: 60-68-178.29) populations compared to non-obesity individuals underscore ET-1’s potent association with adiposity progression and its potential role as a biomarker of vascular and metabolic dysfunction.

These findings align with prior research demonstrating ET-1’s involvement in endothelial dysfunction, vasoconstriction, and pro-inflammatory pathways that contribute to the pathogenesis of obesity-related cardiovascular and renal complications (Kohan et al., 2011; Dhaun et al., 2014). ^17,18^

The dose-dependent increase in ET-1’s effect size from Obesity I to Obesity II suggests a progressive pathophysiological impact, possibly reflecting escalating endothelial injury and metabolic derangement as adiposity worsens. However, the exceptionally high ORs warrant cautious interpretation, as they may reflect residual confounding or threshold effects in ET-1 expression that amplify risk disproportionately in severe obesity. ^19,20^ Concurrently, the TG/HDL-C ratio demonstrated a statistically significant association with obesity severity, with an OR of 2.94 (95% CI: 1.75-4.12) for Obesity II compared to Obesity I. This finding corroborates extensive literature identifying the TG/HDL-C ratio as a reliable surrogate marker of atherogenic dyslipidemia and insulin resistance (Dobiasova 2004; Niroumand et al., 2015). ^21,22,23^

The moderate yet significant increase in odds suggests that lipid abnormalities intensify with advancing obesity, contributing to heightened cardiometabolic risk. Utility in primary healthcare settings, particularly in resource-limited environments. ^29,30,31,32^ The combined evaluation of ET-1 and the TG/HDL-C ratio offers a nuanced understanding of the paradoxical role of adiposity in cardiovascular and metabolic health. ^24^ While traditional metrics such as BMI provide a crude measure of adiposity, these biomarkers capture underlying pathophysiological processes, vascular dysfunction, and lipid metabolism that more accurately reflect individual risk profiles. This biomarker synergy supports a predictive, preventive, and personalized medicine (3PM) approach, enabling early identification of high-risk individuals and tailored intervention strategies. ^25^

## Strengths and limitations

Several limitations merit consideration. The observational design precludes causal inferences, and the potential for residual confounding remains despite multivariate adjustment.

The extreme ORs observed for ET-1 may also indicate population-specific factors or measurement variability. Future longitudinal studies with larger, diverse cohorts are essential to validate these associations and clarify ET-1’s mechanistic role in obesity progression.

## CONCLUSIONS

In conclusion, this study reinforces the value of integrating ET-1 and TG/HDL-C ratio into cardiovascular risk stratification frameworks for patients with ABCD and CKM syndrome. Their robust associations with obesity severity and metabolic risk highlight their potential to transform primary healthcare by enabling early, personalized interventions aligned with national health security and Sustainable Development Goals (SDGs). Such biomarker-driven strategies promise to enhance clinical outcomes and reduce the global burden of non-communicable diseases.

## Data Availability

All data produced in the present study are available upon reasonable request to the authors

## Acknowledgments

The authors gratefully acknowledge the invaluable support and contributions of the clinical and laboratory staff at the participating primary healthcare centers. We extend our sincere appreciation to all study participants for their time and commitment, which made this research possible. This work aligns with national health security priorities and Sustainable Development Goals (SDGs), and we acknowledge the guidance and encouragement from the Ministry of Health and relevant policy stakeholders. Finally, the authors declare no conflicts of interest related to this study.

## Contributors

AAP, AI and SPW conceptualized the study.

### Competing interests

None declared.

### Patient and public involvement

Patients were involved in the design, or conduct, or reporting, or dissemination plans of this research.

### Patient consent for publication

Not applicable.

### Ethics approval

Ethical approval for this study was obtained from the Health Research Ethics Commission General Hospital Dr. Saiful Anwar, Malang, Republic of Indonesia, with ethical approval number 400/055/K.3.102.7/2023. The research was conducted in accordance with the ethical principles outlined in the Declaration of Helsinki (2013). All participants were provided with written and verbal explanations of the study’s purpose, procedures, potential risks, and benefits prior to enrollment.

### Provenance and peer review

Not commissioned; externally peer-reviewed.

### Data availability statement

Data are available upon request. Quantitative data can be made available on reasonable request.

### Open access

This is an open access article distributed in accordance with the Creative Commons Attribution 4.0 Unported (CC BY 4.0) license, which permits others to copy, redistribute, remix, transform and build upon this work for any purpose, provided the original work is properly cited, a link to the licence is given, and indication of whether changes were made. See: https://creativecommons.org/licenses/by/4.0/.

### ORCID iDs

Aldian Arie Pratama

Agustin Iskandar

Singgih Pudjo Wahono

## Notes

### Competing Interest Statement

The authors have declared no competing interest.

### Funding Statement

This study did not receive any funding

### Author Declarations

Ethical approval for this study was obtained from the Health Research Ethics Commission General Hospital Dr. Saiful Anwar, Malang, Republic of Indonesia, with ethical approval number 400/055/K.3.102.7/2023.

